# Additional Roles Reimbursement Scheme commissioning 2020-2023: associations with patient experience and QOF

**DOI:** 10.1101/2024.02.22.24302583

**Authors:** Chris Penfold, Jialan Hong, Peter J Edwards, Mavin Kashyap, Chris Salisbury, Ben Bennett, John Macleod, Maria Theresa Redaniel

## Abstract

**Background:** The Additional Roles Reimbursement Scheme (ARRS) was introduced by NHS England in 2020 alongside Primary Care Networks (PCNs) with aims of increasing the workforce and improving patient outcomes.

**Aim:** Describe the uptake of direct-patient care (DPC)-ARRS roles and its impact on patients’ experiences.

**Design and Setting:** Ecological study using 2020-2023 PCN and Practice workforce data, registered patient characteristics, the General Practice Patient Survey, and the Quality and Outcomes Framework (QOF).

**Methods:** Descriptive statistics with associations examined using quantile and linear regression.

**Results:** By March 2023, 17,588 FTE DPC-ARRS roles were commissioned by 1,223 PCNs. PCNs with fewer constituent practices had more DPC-ARRS roles per population (p<0.001) as did PCNs with more FTE GPs per population (p=0.005). DPC-ARRS commissioning did not vary with age, proportion female or deprivation of practice populations. DPC-ARRS roles were associated with small increases in patient satisfaction (0.8 percentage points increase in patients satisfied per one DPC-ARRS FTE) and perceptions of access (0.7 percentage points increase in patients reporting ‘good’ experience of making an appointment per one DPC-ARRS FTE), but not with overall QOF achievement.

**Conclusions:** The commissioning of DPC-ARRS roles was associated with small increases in patient satisfaction and perceptions of access, but not with QOF achievement. DPC-ARRS roles were employed in areas with more GPs rather than compensating for a shortage of doctors. Single practice PCNs commissioned more roles per registered population, which may be advantageous to single practice PCNs. Further evaluation of the scheme is warranted.

**How this fits in:**

1. Primary Care Networks (PCNs) commissioning of non-GP direct-patient care (DPC) roles via the Additional Roles Reimbursement Scheme (ARRS) has expanded rapidly, with an allocated budget of £110m in 2019/2020 employing 279 full time equivalent (FTE) DPC-ARRS staff in March 2020, to £1,027m in 2022/2023, employing 17,588 FTE DPC-ARRS staff in March 2023.
2. Previous research, using NHS England data prior to 2020, had reported associations between increased Healthcare Associate Professionals employment and reductions in patient satisfaction and perceptions of access, with no impact on Quality and Outcomes Framework (QOF) achievement, but it was not known if these trends remained after the implementation of ARRS.
3. This study found a small increase in both overall patient satisfaction and perceptions of access, which equates to approximately 240-400 patients satisfied with their care and 210-350 patients able to make appointments for each FTE in ARRS roles employed in a typical PCN (30,000-50,000 patients), but no association with overall QOF achievement.
4. Further evaluation is required to identify if the observed associations can be attributed to the ARRS roll-out and if this represents value for money.

## Introduction

The primary care workforce challenge in England is complex and long-standing, with shortfalls of General Practitioners (GPs) and practice nurses alongside increasing healthcare demands (1). In 2019, NHS England established Primary Care Networks (PCNs) to increase the primary care workforce in order to improve personalised and integrated care (2). Commissioned by PCNs, the Additional Roles Reimbursement Scheme (ARRS) expands the role of non-medical practitioners in primary care (3) to improve care delivery, expedite patient access, mitigate rising demand, and provide an advanced career pathway for non-GP practitioners (4,5). Eligible ARRS roles initially included: social prescribing link workers, clinical pharmacists, first contact physiotherapists, physician associates, and paramedics (3). The eligible roles have been expanded annually to include other direct patient care (DPC) and administrative roles (6), and the ARRS budget was £1,027m in 2022/2023 (7).

The broadening of the skill mix in NHS primary care through the ARRS is occurring rapidly (8). Evaluations of the scheme have focussed on health inequalities. Qualitative research identified that the ARRS scheme may exacerbate inequalities through recruitment challenges for areas of high deprivation and their inability to compete financially with wealthier PCNs (9). Whereas analysis of NHS primary care workforce data found that the introduction of PCNs and the overall commissioning of roles through the ARRS did not exacerbate existing inequalities in clinical staff distribution (10).

Previously, the introduction of new roles in NHS primary care in England was associated with worse patient satisfaction, increased health service costs, and no effect on overall Quality and Outcomes Framework (QOF) achievement, an indicator of clinical effectiveness (11,12). However, broadening the skill mix helped tackle workforce shortages and free GP time for complex patients (12–14).

The aims of this study are to describe variation in the commissioning of ARRS roles during the first three years of the scheme and to determine the impact of this initial phase of the scheme on patients’ experiences of primary care services and on clinical effectiveness. This will inform the implementation of the ARRS and future schemes to broaden the primary care skill mix.

## Methods

### Study design

An ecological study with outcomes recorded at PCN-level (role commissioning) and Practice-level (patient experiences, clinical effectiveness). We included Practices and PCNs in the 2020-2023 PCN and GP Workforce datasets, and the 2023 General Practice Patient Survey (GPPS). We excluded Practices with no registered patients and those not in PCNs since they cannot commission roles through the ARRS.

### Data sources

We used openly accessible data covering 2020-2023 from the following sources (full details of data sources and quality in the Supplementary material):

#### 1. PCN workforce

Quarterly PCN-level full-time equivalent (FTE) employment for 15 DPC staff roles (as of March 2023) funded through the ARRS scheme (15), see Table S1 for a full list of included and excluded ARRS roles.

#### 2. General Practice workforce

Quarterly snapshots of FTE for NHS primary care GPs, nurses, DPC, and administrative staff (16).

#### 3. General Practice Patient Survey (GPPS)

The GPPS, run annually, includes responses from >700,000 patients aged ≥16 years about their experiences of General Practices (17). GPPS fieldwork takes place between January and March/April each year.

#### 4. Quality and Outcomes Framework (QOF)

The QOF is a pay-for-performance scheme to assess and reward the quality of care provided by Practices (18,19). Results are published annually.

#### 5. General Practice registered and weighted population (1st April 2021, NHS Digital)

Quarterly publication of the number of patients registered at each General Practice in England from each of the smallest administrative geographical areas (Lower-layer Super Output Areas, LSOA) (20). Payments to General Practices and PCNs are adjusted to create a “weighted patient count” using the Carr-Hill formula to reflect the perceived need of the registered population.

#### 6. Indices of Multiple Deprivation (IMD)

The English Indices of Multiple Deprivation measure relative levels of deprivation in the 32,844 LSOAs in England (21).

### Exposure

Our exposure was the FTE of the DPC-ARRS roles at Practice-level. The PCN workforce data does not record how ARRS roles are shared across Practices within PCNs. We assumed an equal allocation of the FTE across Practices, e.g. that one FTE paramedic in a four Practice PCN was shared equally as 0.25 FTE per Practice.

### Outcomes

#### Patient experience

Measured as the proportion of patients able to access care and proportion satisfied with their care (11). These were derived from two GPPS questions relating to (i) patient’s experience of making an appointment (access), and, (ii) their overall experience (satisfaction), using a five-point Likert scale. The responses ‘very good’ and ‘fairly good’ were combined as positive indicators of access or satisfaction. GPPS fieldwork is done January-April each year so we used the March 2023 PCN workforce to most closely reflect the workforce experienced by GPPS respondents.

#### Clinical effectiveness

We used the total QOF points achieved across all domains as a proportion of the maximum available QOF points, captured as a percentage, to indicate clinical effectiveness.

### Covariates

Covariates included were the unweighted number of registered patients, the demographics of the registered Practice population (mean age, proportion female, area-level deprivation), and the FTE of GPs and nurses.

#### Estimation of Practice and PCN-level deprivation

The population weighted mean IMD rank of Practices was calculated as the sum of the IMDs of the LSOAs of registered patients divided by the proportion of the Practice population from that LSOA. For PCNs we aggregated Practices within PCNs.

### Statistical analysis

#### Temporal trends of ARRS roles

We described the cumulative commissioning of ARRS roles per quarter from March 2020 to March 2023. We described how the number of patients, weighted by the Carr-Hill formula, per FTE ARRS roles in March 2023 varied by quintiles of PCN-level characteristics: mean age of patients, number of patients per FTE GP, proportion female, area-level deprivation, and the number of Practices in the PCN. We used quantile regression to test for trends.

#### Association of ARRS FTE with patient experience and clinical effectiveness

We used linear regression models to estimate associations of ARRS FTE with patient satisfaction, access to care, and QOF achievement. Models were adjusted for the unweighted number of registered patients and weighted by the number of respondents to the relevant GPPS question (patient experience outcomes). Remaining covariates were included as further adjustments. We included two-way interactions between ARRS FTE and GP FTE and nurse FTE to determine the complementarity of ARRS roles with GPs and nurses.

We used R version 4.2 (22) and the ‘tidyverse’ (23) and ‘quantreg’ packages (24).

#### Sensitivity analyses

1. Regression models for patient outcomes may indicate reverse causality. We repeated our fully adjusted regression models with 2020 GPPS outcomes to account for this potential effect.
2. We assumed ARRS FTE was allocated equally across Practices within PCNs. We repeated analyses but allocated ARRS FTE by unweighted registered practice population.

## Results

We included 1,253 PCNs and 6,771 GP Practices who have submitted data to NHS Digital March 2020. Seventy-six Practices (1.1%) were not part of a PCN and excluded from our study.

### PCN commissioning of ARRS roles

In March 2020, 158 of 1,253 PCNs (12.6%) reported commissioning a total of 279 FTE staff DPC ARRS roles (Figure 1). By March 2023, 1,223 of 1,263 PCNs (96.8%) had commissioned 17,588 FTE. The main roles commissioned according to FTE were pharmacists (4,783 FTE March 2023), pharmacy technicians (1,460 FTE), care coordinators (3,217 FTE), social prescribing link workers (2,635 FTE), and physiotherapists (1,393 FTE, Figure 2 and Table S2). The median PCN-level FTE of ARRS staff roles was 12.6 (IQR: 8.8, 17.9) in March 2023 and a median of 2.6 FTE ARRS roles per Practice (IQR: 1.7, 3.8).

**Figure 1:**
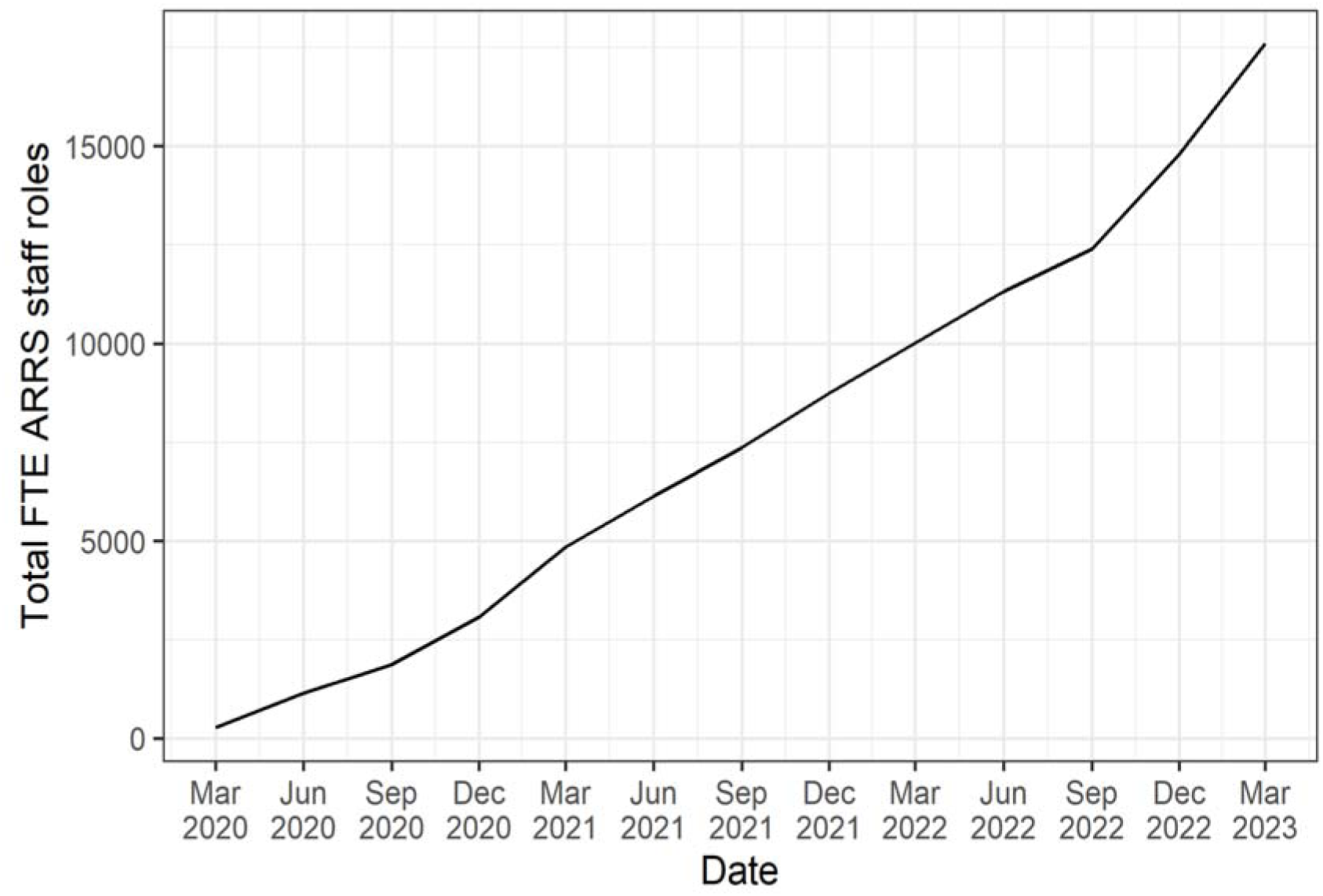
The total FTE of ARRS funded staff roles in England between March 2020 and March 2023.

**Figure 2:**
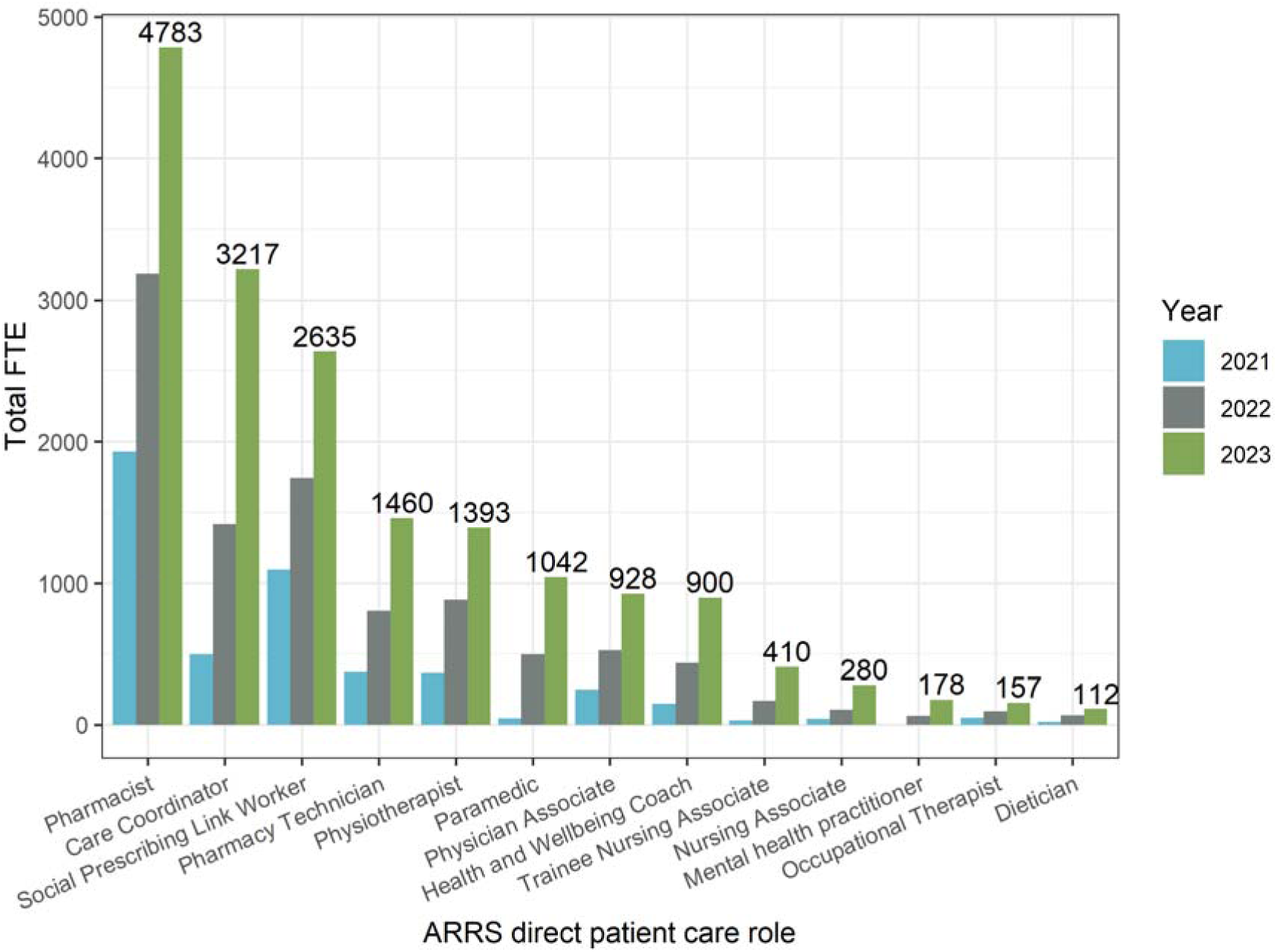
FTE in ARRS DPC roles annually 2021-2023, by role. Note: Roles with <100 FTE in 2023 have been excluded from this plot for clarity. These were (March 2023 FTE): Advanced Nurse Practitioner (47), Podiatrist (45)

### Variation in ARRS commissioning by practice characteristics

In March 2023 the median PCN-level FTE ARRS roles per 10,000 registered patients (weighted by the Carr-Hill formula) was 2.91 (IQR: 2.06-3.80). Figure 3 highlights the variation in the commissioning of ARRS roles by the characteristics of the PCN population. PCNs with the most GP FTE per 10k_weighted_ patients had around 0.4 more FTE ARRS roles compared with those with the least GP FTE (2.73 versus 3.11 least versus most GP FTE, p_trend_=0.005). The number of Practices within PCNs was negatively associated with the FTE in ARRS roles per patient. PCNs comprised of one Practice compared with the quintile of PCNs comprised of the most Practices (7 to 22) had nearly 0.6 more FTE in ARRS roles per 10k patients (3.51 versus 2.95 FTE ARRS/10k_weighted_ patients, p_trend_=0.001). The FTE in ARRS roles per 10k patients varied minimally by mean age of patients (median 2.81 versus 3.00 youngest versus oldest, p_trend_=0.330), proportion female (2.89 versus 2.98 smallest versus largest proportion, p_trend_=0.231), or their area-level deprivation (2.92 versus 2.98 most versus least deprived, p_trend_=0.603).

**Figure 3:**
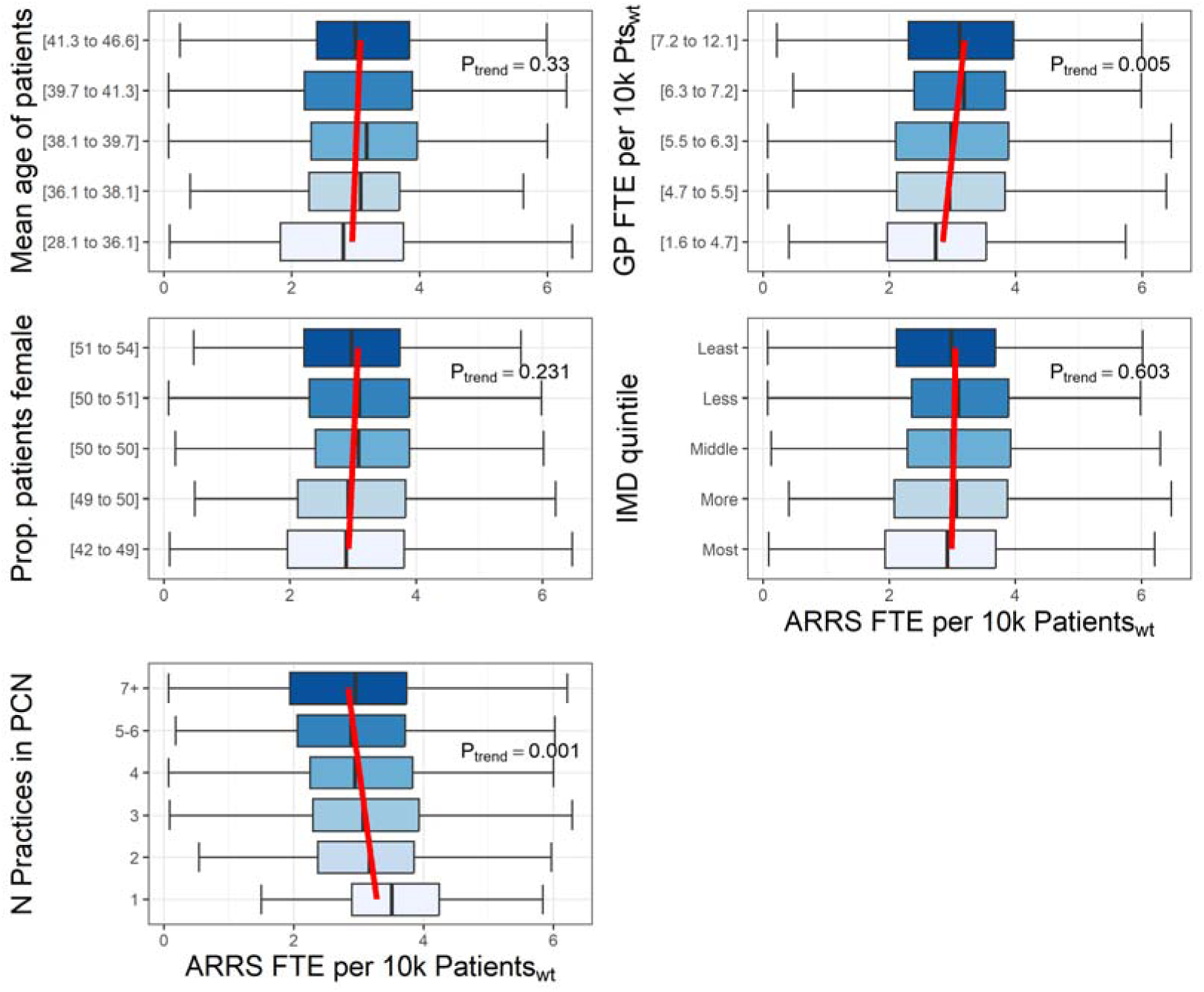
The variation in the PCN-level median ARRS FTE per weighted number of patients by registered patient age, proportion female, area-level deprivation of registered patients, GP FTE per weighted number of patients, and number of Practices per PCN Note – red lines represent the linear trend of median ARRS FTE per patient

**Figure 4:**
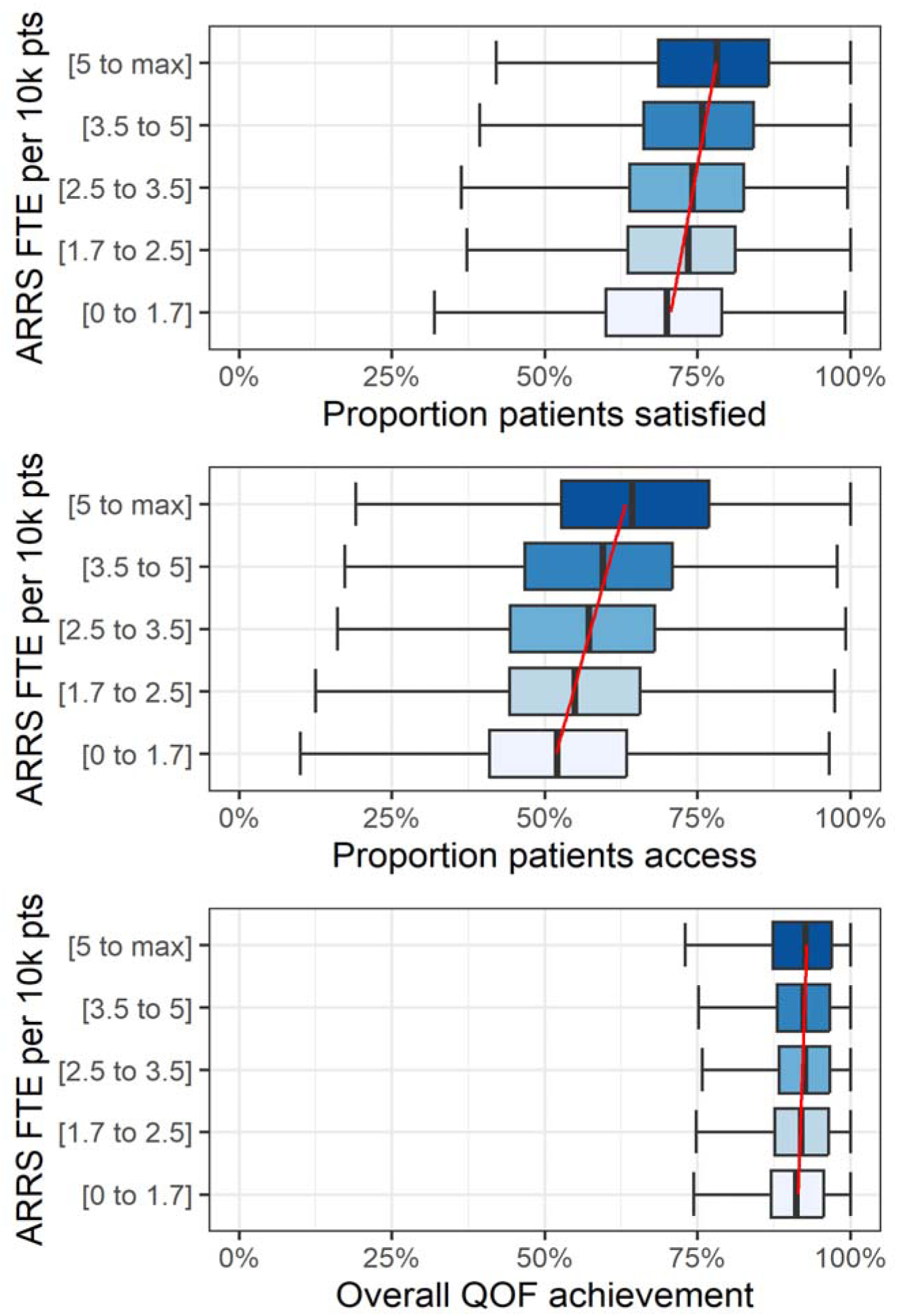
Box plots of the Practice-level proportion of patients satisfied with their GP service and able to make an appointment (access), and overall QOF achievement percentage by quintiles of ARRS FTE per 10,000 patients Note – red lines represent the linear trend from quantile regression across exposure categories of median outcome values

### Patient outcomes

In March 2023 the overall proportion of patients who reported a ‘good’ experience making an appointment at their GP was 54.4% and the proportion satisfied with GP services was 71.3%. Practices with more ARRS FTE per 10k patients had a higher proportion of patients satisfied and able to make appointments (Figure 3). The percentage of overall QOF achievement varied minimally by ARRS FTE. Practices with the most ARRS FTE achieved 1.5 percentage points more of their QOF overall achievement percentage compared to Practices with the least ARRS FTE.

In our adjusted regression models an increase of one FTE in ARRS roles was associated with a 0.8 percentage point increase (95% CI: 0.6%-1.0%, p<0.001) in the proportion of patients satisfied with their care and a 0.7 percentage point increase (95% CI: 0.5%-1.0%, p<0.001) in the proportion of patients able to make appointments (Table 1). This equates to an increase of approximately 240-400 patients satisfied with their care and 210-350 patients able to make appointments for each FTE in ARRS roles employed in a typical PCN (30,000-50,000 patients). Whereas for overall QOF achievements a one FTE increase in ARRS roles was associated with a 0.04 percentage point decrease in percentage overall achievement, but with confidence intervals extending above the null value (95% CI: −0.21% to 0.12%, p=0.6).

**Table 1:**
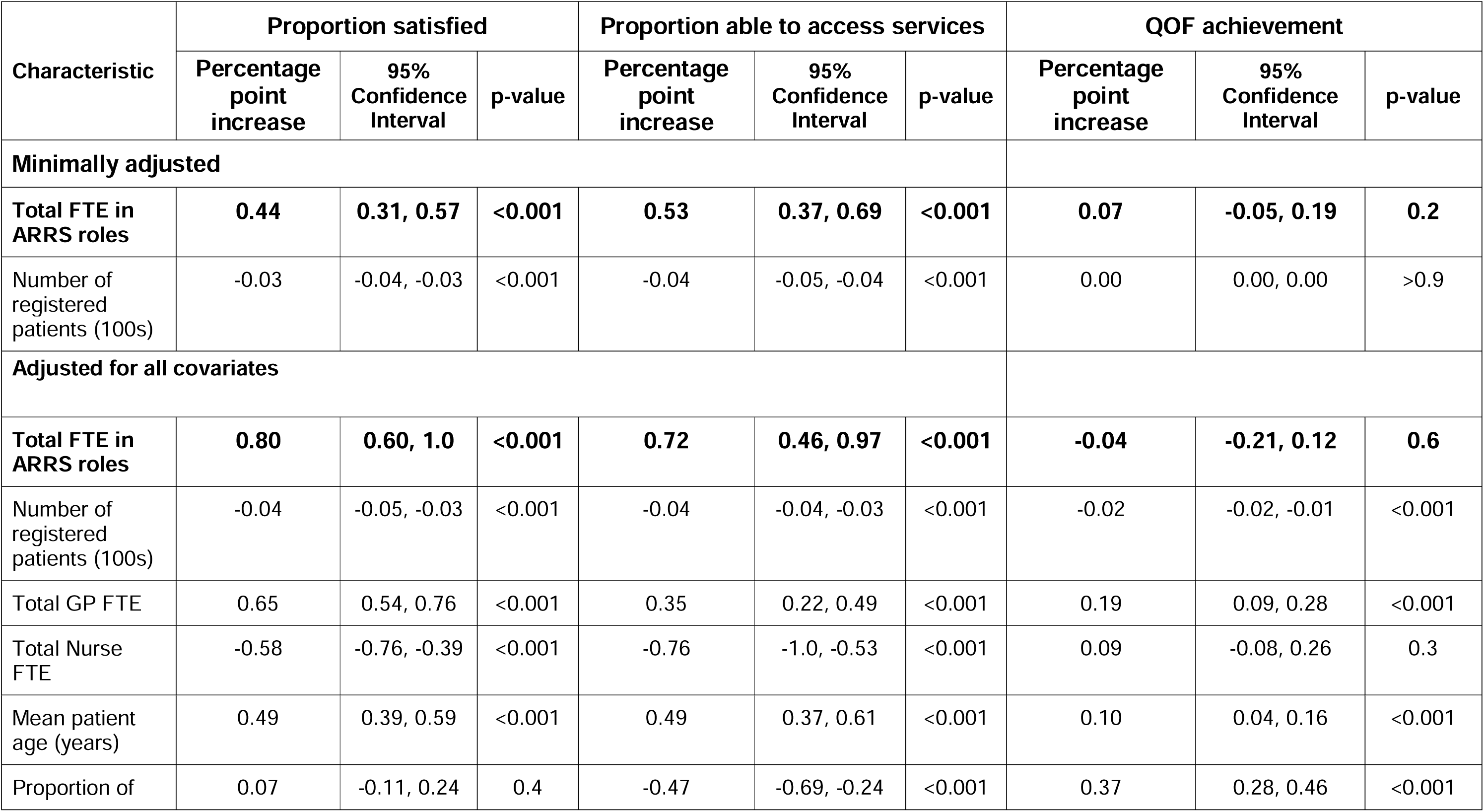
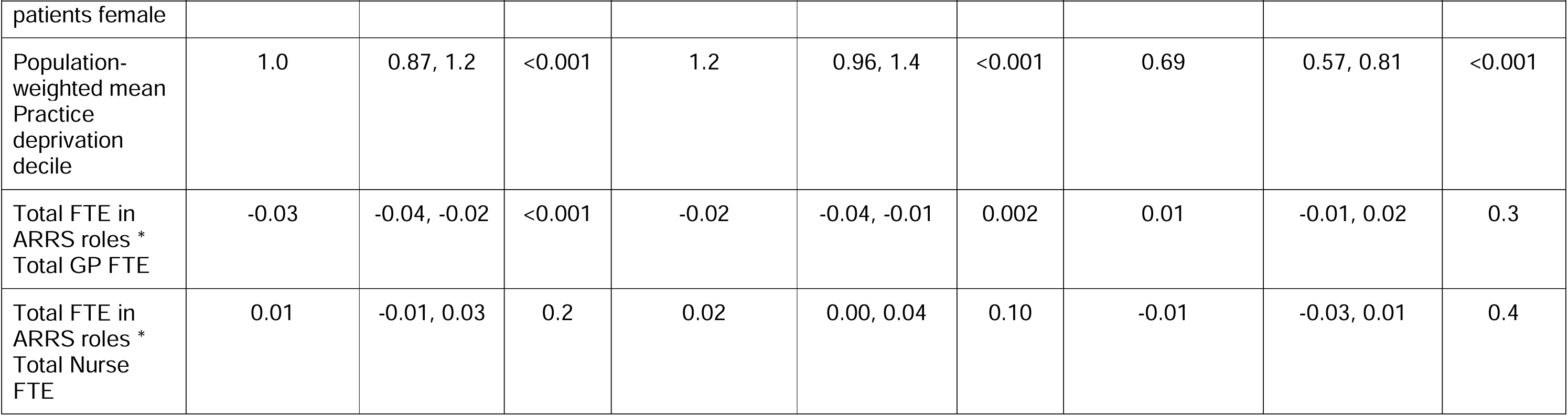
Linear regression models of ARRS FTE against the proportion of GPPS respondents satisfied with and able to access primary medical care, and QOF overall achievement percentage.

#### Sensitivity analysis 1 – adjustment for 2020 GPPS outcomes

Inclusion of the 2020 GPPS outcomes for satisfaction and access reduced the effect sizes but the confidence intervals still supported the observed positive associations (Table S3).

#### Sensitivity analysis 2 – Practice population weighted deployment of ARRS roles

Changing our assumption about how ARRS roles were deployed across Practices within PCNs from an equal distribution to a registered population weighted approach reduced the effect size estimates of our regression models, but they still supported the primary analyses (Table S4).

## Discussion

### Summary

The introduction of the ARRS has, by March 2023, increased the number of staff in direct patient care roles by more than 17,000 FTE. The commissioning of DPC-ARRS roles does not vary by area-level deprivation. DPC-ARRS roles were employed in areas with more GPs rather than compensating for a shortage of doctors. Single Practice PCNs commissioned more FTE DPC-ARRS roles than PCNs with multiple Practices.

### Strengths and weaknesses

The main strengths are that the findings are representative of the whole of England and that we linked openly available public data. PCN workforce data was initially quite incomplete, but improved rapidly and are now likely to be accurate and reliable. Also, that we included patient outcomes, the longitudinal nature of the PCN workforce data, and that the patient outcomes we used from the GPPS have been used in previous studies (11,12) which facilitates comparisons.

The ecological study design is an important weakness. We also do not know how PCNs deployed their ARRS workforce. We explored two alternative assumptions and findings were consistent. Early in the ARRS the completeness of workforce reporting by PCNs was relatively poor. We could not differentiate between PCNs not commissioning any ARRS roles and low engagement with workforce reporting.

### Comparison with existing literature

We found no variation in the commissioning of DPC-ARRS roles by area-level deprivation, in agreement with previous analysis of PCN workforce data (10), but opposite of qualitative interviews with NHS staff (9). These discrepant findings may be because PCNs in more deprived areas are commissioning staff through the scheme, but not the staff they want or need for their population’s needs. Alternatively, as reported elsewhere (25–27), the Carr-Hill adjustment does not sufficiently account for the additional health needs of more deprived populations. A Health Foundation report found that by using the new NHS England PCN-adjusted population rather than the Carr-Hill adjustment, PCNs in more deprived areas had fewer ARRS staff than those in less deprived areas (28).

PCNs comprised of fewer Practices commissioned more ARRS roles than those with more Practices, and was most pronounced for single Practice PCNs. A possible explanation for our finding is that PCNs with fewer Practices are organisationally simpler, meaning that employing staff through ARRS is more akin to employing staff directly through the Practice. This could incentivise Practice mergers, which may be at the expense of patient satisfaction and access, and continuity of care, which are reduced in large Practices (29,30). It has been highlighted previously that >40% PCNs were not of the recommended size and that this may affect their ability to effectively utilise investment (31). Our findings support this concern.

Commissioning of ARRS roles followed similar trends to that of GPs. PCNs with more GP FTE per 10k needs adjusted patients commissioned more DPC-ARRS FTE. Whether this addresses the needs of the local population or is a further reflection of local staff recruitment challenges is unknown. The potential for this trend to exacerbate existing inequalities in the distribution of the primary care workforce does not align with other research, which found this was not the case (10).

Practices with more FTE in ARRS roles had slightly better patient reported satisfaction and ability to make appointments, but the effect size was very small, especially given the investment in the scheme. This is the opposite of cross-sectional (12) and longitudinal (11) findings using the same GPPS outcomes in 2019 and earlier. These studies were conducted during a period of relative stability before the COVID-19 pandemic whereas our study may have been affected by restrictions imposed during the pandemic, despite our primary end point being in the post-pandemic recovery period. Additionally, the transfer of tasks from GPs to non-GP practitioners was identified as a key challenge to be faced by the newly formed PCNs (32). Our outcome was three years after PCNs were introduced, which may have given PCNs sufficient time to address this challenge.

### Implications for research and practice

The ARRS scheme expanded existing roles and introduced new roles into NHS primary care in England. It remains unclear if minimal increases in patient perceptions of access and overall care represent value for money. The relationship between PCN organisational structure and commissioning of ARRS roles may have implications for the evolution of PCNs.

Future research could address the lack of patient-level data on consultations with ARRS staff. This would give a better understanding of the types of patients seen by each of the roles and outcomes of those consultations, which would inform overall workforce planning, training needs of staff in these roles, and opportunities to develop these roles. Finally, it may be beneficial to understand how and why the structure of PCNs affects their ability to commission ARRS roles.

## Data Availability

All data are publicly available

## Ethical approval

Ethical approval for this study was not required.

## Acknowledgements

No acknowledgments

