## Supplementary material for "Additional Roles Reimbursement Scheme commissioning 2020-2023: associations with patient experience and QOF"

### Data sources

Further details of the openly accessible data we used openly accessible data covering 2020 to 2023, unless stated otherwise:

1. PCN workforce – NHS England

Quarterly PCN-level full-time equivalent (FTE) employment for 15 DPC staff roles funded through the ARRS scheme. This includes Advanced Practitioner (AP) roles, which were combined with their non-AP equivalents where relevant. Collection of workforce data at PCN-level started in February 2020 through the National Workforce Reporting Service (NWRS). Engagement with the NWRS varied substantially between PCNs after its introduction meaning that valid data were available for only approximately 15% PCNs in March 2020, rising to around 96% by March 2023. The PCN workforce data does not provide reliable estimates of staff headcount, but weekly working hours are reliable and are used to estimate proxy FTE.

2. General Practice workforce – NHS England

Quarterly (until July 2021) and monthly (from August 2021) snapshots of FTE for NHS primary care GPs, nurses, DPC, and administrative staff. Approximately 99.5% of Practices submitted valid data for each reporting period from 2019 onwards, although coverage may have been slightly lower during the early months of the COVID-19 pandemic.

3. General Practice Patient Survey (GPPS)

The GPPS, run annually by IPSOS on behalf of NHS England, includes responses from >700,000 patients aged 16 years and over to assess their experiences of primary healthcare services delivered by General Practices. Fieldwork for the GPPS typically takes place between January and March/April each year.

4. Quality and Outcomes Framework (QOF)

The QOF covers all General Practices in England and is a pay-for-performance scheme intended to assess and reward the quality of care provided by Practices (18). The QOF assigns clinical and organizational indicators to practices, focusing on areas such as clinical effectiveness and patient experience. Practices earn points based on performance, with financial incentives linked to these points. The 2022/2023 QOF is comprised of five domains and overall domain achievement.

5. General Practice registered and weighted population

Quarterly publication of the number of patients registered at each General Practice in England from each of the smallest administrative geographical areas (Lower-layer Super Output Areas, LSOA) in England (20). Payments to General Practices and PCNs reflect the perceived need of the registered population. Each Practice’s registered patient list is adjusted according to the Carr-Hill Formula to take into consideration differences in the age and sex of the patients as well as any in nursing or residential care, additional patient need due to medical conditions, patient turnover and unavoidable costs based upon rurality and staff market forces for the area. This results in an adjusted count of patients known as the “weighted patient count”.

6. Indices of Multiple Deprivation (IMD)

The English Indices of Multiple Deprivation measure relative levels of deprivation in the 32,844 LSOAs in England.

#### Table S1: Roles eligible for ARRS funding in March 2023 and their inclusion/exclusion as direct patient care roles

| **ARRS roles 2023** | **Grouped role** | **Direct patient care role** |
| --- | --- | --- |
| Advanced Dietician Practitioner | Dietician | Yes |
| Dietician | Dietician | Yes |
| Advanced Nurse Practitioner |  | Yes |
| Advanced Occupational Therapist Practitioner | Occupational Therapist | Yes |
| Therapist- Occupational Therapist | Occupational Therapist | Yes |
| Advanced Paramedic Practitioner | Paramedic | Yes |
| Paramedic | Paramedic | Yes |
| Advanced Pharmacist Practitioner | Pharmacist | Yes |
| Pharmacist | Pharmacist | Yes |
| Advanced Physiotherapist Practitioner | Physiotherapist | Yes |
| First Contact Physiotherapist | Physiotherapist | Yes |
| Advanced Podiatrist Practitioner | Podiatrist | Yes |
| Podiatrist | Podiatrist | Yes |
| Care Coordinator |  | Yes |
| Health Support Worker | Mental health practitioner | Yes |
| Health and Wellbeing Coach |  | Yes |
| High Intensity Therapist | Mental health practitioner | Yes |
| Nursing Associate |  | Yes |
| Pharmacy Technician |  | Yes |
| Physician Associate |  | Yes |
| Psychological Wellbeing Practitioner | Mental health practitioner | Yes |
| Social Prescribing Link Worker |  | Yes |
| Therapist- Other | Mental health practitioner | Yes |
| Trainee Nursing Associate |  | Yes |
| Trainee Psychological Wellbeing Practitioner | Mental health practitioner | Yes |
| General practice assistant |  | No |
| Digital and transformation lead |  | No |

#### Table S2: The total FTE in ARRS funded staff roles in England between March 2020 and March 2023, by role type

| **Staff role** | **2020** | | | | **2021** | | | | **2022** | | | | **2023** |
| --- | --- | --- | --- | --- | --- | --- | --- | --- | --- | --- | --- | --- | --- |
|  | **Mar** | **Jun** | **Sep** | **Dec** | **Mar** | **Jun** | **Sep** | **Dec** | **Mar** | **Jun** | **Sep** | **Dec** | **Mar** |
| Pharmacist | 153 | 609 | 947 | 1,375 | 1,929 | 2,275 | 2,626 | 2,923 | 3,183 | 3,451 | 3,684 | 4,209 | 4,783 |
| Care Coordinator | 0 | 0 | 0 | 132 | 498 | 682 | 888 | 1,108 | 1,418 | 1,682 | 1,895 | 2,448 | 3,217 |
| Social Prescribing Link Worker | 112 | 431 | 616 | 852 | 1,093 | 1,261 | 1,427 | 1,612 | 1,741 | 1,903 | 2,047 | 2,345 | 2,635 |
| Pharmacy Technician | 3 | 24 | 95 | 215 | 373 | 489 | 562 | 682 | 807 | 927 | 1,048 | 1,246 | 1,460 |
| Physiotherapist | 6 | 47 | 134 | 220 | 364 | 508 | 652 | 779 | 884 | 977 | 1,032 | 1,178 | 1,393 |
| Paramedic | 2 | 4 | 12 | 25 | 47 | 160 | 256 | 388 | 498 | 612 | 697 | 869 | 1,042 |
| Physician Associate | 2 | 36 | 65 | 161 | 249 | 332 | 395 | 460 | 527 | 607 | 657 | 781 | 928 |
| Health and Wellbeing Coach | 0 | 0 | 0 | 60 | 148 | 198 | 252 | 357 | 438 | 510 | 578 | 736 | 900 |
| Trainee Nursing Associate | 0 | 0 | 0 | 8 | 31 | 41 | 69 | 119 | 169 | 220 | 250 | 325 | 410 |
| Nursing Associate | 0 | 0 | 0 | 6 | 41 | 56 | 73 | 88 | 107 | 137 | 153 | 206 | 280 |
| Mental health practitioner | 0 | 0 | 0 | 0 | 0 | 41 | 54 | 69 | 65 | 86 | 113 | 144 | 178 |
| Occupational Therapist | 0 | 2 | 10 | 17 | 49 | 57 | 72 | 85 | 95 | 109 | 123 | 145 | 157 |
| Dietician | 0 | 0 | 1 | 8 | 21 | 28 | 38 | 51 | 67 | 80 | 85 | 97 | 112 |
| Advanced Nurse Practitioner | 0 | 0 | 0 | 0 | 0 | 0 | 0 | 0 | 0 | 0 | 0 | 20 | 47 |
| Podiatrist | 0 | 0 | 1 | 3 | 7 | 12 | 14 | 20 | 22 | 26 | 33 | 44 | 45 |
| **Total** | **279** | **1,154** | **1,880** | **3,083** | **4,851** | **6,139** | **7,380** | **8,739** | **10,021** | **11,327** | **12,395** | **14,794** | **17,588** |

#### Table S3: Inclusion of 2020 GPPS outcomes in linear regression models of ARRS FTE against the proportion of GPPS respondents satisfied with and able to access primary medical care, weighted by number of GPPS question respondents

| **Characteristic** | **Proportion satisfied** | | | **Proportion able to access services** | | |
| --- | --- | --- | --- | --- | --- | --- |
|  | **Adjusted for all covariates** | | | **Adjusted for all covariates** | | |
|  | **Beta^1^** | **95% CI**^2^ | **p-value** | **Beta^1^** | **95% CI**^2^ | **p-value** |
| **Total FTE in ARRS roles** | **0.57** | **0.40, 0.73** | **<0.001** | **0.32** | **0.12, 0.52** | **0.002** |
| Proportion satisfied (2020 GPPS) | 0.75 | 0.73, 0.78 | <0.001 |  |  |  |
| Proportion able to access (2020 GPPS) |  |  |  | 0.68 | 0.66, 0.70 | <0.001 |
| Number of registered patients (100s) | -0.02 | -0.03, -0.02 | <0.001 | -0.02 | -0.02, -0.01 | <0.001 |
| Total GP FTE | 0.28 | 0.19, 0.37 | <0.001 | 0.14 | 0.03, 0.24 | 0.014 |
| Total Nurse FTE | -0.31 | -0.46, -0.15 | <0.001 | -0.38 | -0.57, -0.20 | <0.001 |
| Mean patient age (years) | 0.19 | 0.11, 0.27 | <0.001 | 0.22 | 0.13, 0.32 | <0.001 |
| Proportion of patients female | -0.12 | -0.26, 0.03 | 0.12 | -0.25 | -0.42, -0.07 | 0.006 |
| Population-weighted mean Practice deprivation decile | 0.31 | 0.16, 0.47 | <0.001 | 0.28 | 0.10, 0.46 | 0.002 |
| Total FTE in ARRS roles * Total GP FTE | -0.02 | -0.03, -0.01 | <0.001 | -0.01 | -0.02, 0.00 | 0.066 |
| Total FTE in ARRS roles * Total Nurse FTE | 0.02 | 0.00, 0.03 | 0.035 | 0.02 | 0.01, 0.04 | 0.007 |

^1^Percentage point increase/decrease in proportion satisfied/access

^2^CI = Confidence Interval

#### Table S4: Linear regression models of ARRS FTE (Practice population weighted deployment) against the proportion of GPPS respondents satisfied with and able to access primary medical care, and QOF overall achievement percentage

| **Characteristic** | **Proportion satisfied** | | | **Proportion able to access services** | | | **QOF achievement** | | |
| --- | --- | --- | --- | --- | --- | --- | --- | --- | --- |
|  | **Adjusted for practice population** | | | **Adjusted for practice population** | | | **Adjusted for practice population** | | |
|  | Percentage point increase | **95% Confidence Interval** | **p-value** | Percentage point increase | **95% Confidence Interval** | **p-value** | Percentage point increase | **95% Confidence Interval** | **p-value** |
| **Total FTE in ARRS roles** | **0.53** | **0.37, 0.69** | **<0.001** | **0.41** | **0.21, 0.61** | **<0.001** | **0.14** | **-0.02, 0.30** | **0.092** |
| Number of registered patients (100s) | -0.05 | -0.06, -0.04 | <0.001 | -0.05 | -0.06, -0.04 | <0.001 | -0.02 | -0.03, -0.01 | <0.001 |
| Total GP FTE | 0.51 | 0.40, 0.62 | <0.001 | 0.17 | 0.03, 0.30 | 0.014 | 0.21 | 0.12, 0.30 | <0.001 |
| Total Nurse FTE | -0.35 | -0.57, -0.14 | 0.001 | -0.47 | -0.73, -0.20 | <0.001 | 0.14 | -0.03, 0.32 | 0.11 |
| Mean patient age (years) | 0.45 | 0.36, 0.55 | <0.001 | 0.44 | 0.32, 0.57 | <0.001 | 0.10 | 0.03, 0.16 | 0.002 |
| Proportion of patients female | 0.09 | -0.09, 0.26 | 0.3 | -0.44 | -0.66, -0.22 | <0.001 | 0.36 | 0.27, 0.45 | <0.001 |
| Population-weighted mean Practice deprivation decile | 1.2 | 0.97, 1.3 | <0.001 | 1.3 | 1.1, 1.6 | <0.001 | 0.69 | 0.57, 0.81 | <0.001 |
| Total FTE in ARRS roles * Total GP FTE | 0.00 | -0.01, 0.01 | 0.4 | 0.01 | 0.00, 0.02 | 0.2 | 0.00 | -0.01, 0.01 | 0.8 |
| Total FTE in ARRS roles * Total Nurse FTE | -0.01 | -0.02, 0.01 | 0.5 | 0.00 | -0.02, 0.02 | 0.7 | -0.02 | -0.03, 0.00 | 0.10 |
